## Supplementary material for "Predicting willingness to be vaccinated for Covid-19: evidence from New Zealand": S1 Questionnaire

**Covid Vaccination Questionnaire**

SAMPLE: 1000 households nationwide

**Q1: Beliefs about Covid-19**

We are interested in your opinions about Covid-19. How strongly do you agree or disagree with the following statements?

| **Item** | **Strongly agree** | **Agree** | **Unsure/**  **neutral** | **Disagree** | **Strongly disagree** |
| --- | --- | --- | --- | --- | --- |
| You cannot catch Covid-19 from people with the virus who do not have symptoms | ☐ | ☐ | ☐ | ☐ | ☐ |
| Covid-19 is only a danger to the elderly and people who already have health problem | ☐ | ☐ | ☐ | ☐ | ☐ |
| Infected people spread Covid-19 by coughing and sneezing | ☐ | ☐ | ☐ | ☐ | ☐ |
| Children cannot catch Covid-19 | ☐ | ☐ | ☐ | ☐ | ☐ |
| Once you have had Covid-19 you are immune to re-infection | ☐ | ☐ | ☐ | ☐ | ☐ |
| I think Covid-19 is a hoax | ☐ | ☐ | ☐ | ☐ | ☐ |
| Fears about Covid-19 are exaggerated | ☐ | ☐ | ☐ | ☐ | ☐ |
| Covid-19 most likely comes from bats | ☐ | ☐ | ☐ | ☐ | ☐ |
| Covid-19 is a man-made virus | ☐ | ☐ | ☐ | ☐ | ☐ |
| Children are perfectly safe from Covid-19 | ☐ | ☐ | ☐ | ☐ | ☐ |
| You can catch Covid-19 by touching anything handled by an infected person | ☐ | ☐ | ☐ | ☐ | ☐ |
| Covid-19 is no worse than the seasonal flu | ☐ | ☐ | ☐ | ☐ | ☐ |

**Q2: Beliefs about eliminating Covid-19**

We are interested in your opinions about eliminating Covid-19. How strongly do you agree or disagree with the following statements?

| **Item** | **Strongly agree** | **Agree** | **Unsure/**  **neutral** | **Disagree** | **Strongly disagree** |
| --- | --- | --- | --- | --- | --- |
| We need to eliminate Covid-19 from New Zealand to save lives | ☐ | ☐ | ☐ | ☐ | ☐ |
| We should just live with it until we have a vaccine | ☐ | ☐ | ☐ | ☐ | ☐ |
| It would be better to let it spread and build herd immunity | ☐ | ☐ | ☐ | ☐ | ☐ |
| There is no point trying to eliminate Covid-19 because it is a virus and will keep changing | ☐ | ☐ | ☐ | ☐ | ☐ |
| Covid-19 is everywhere in the world so there is no way we can keep it out | ☐ | ☐ | ☐ | ☐ | ☐ |

**Q3: Taking responsibility and action**

We are interested in how strongly you feel about the need for action to be taken to eliminate Covid-19 from New Zealand. How strongly do you agree or disagree with the following statements?

| **Item** | **Strongly agree** | **Agree** | **Unsure/**  **neutral** | **Disagree** | **Strongly disagree** |
| --- | --- | --- | --- | --- | --- |
| Eliminating Covid-19 from New Zealand is the right thing to do | ☐ | ☐ | ☐ | ☐ | ☐ |
| I feel some responsibility for eliminating Covid-19 from New Zealand | ☐ | ☐ | ☐ | ☐ | ☐ |
| I am prepared to change my normal behaviour to eliminate Covid-19 from New Zealand | ☐ | ☐ | ☐ | ☐ | ☐ |
| It is important to work together to eliminate Covid-19 from New Zealand | ☐ | ☐ | ☐ | ☐ | ☐ |
| Nearly everyone I know thinks eliminating Covid-19 from New Zealand is the right thing to do | ☐ | ☐ | ☐ | ☐ | ☐ |
| Most people I know feel some responsibility for eliminating Covid-19 from New Zealand | ☐ | ☐ | ☐ | ☐ | ☐ |
| I think nearly everyone is prepared to change their normal behaviour to eliminate Covid-19 from New Zealand | ☐ | ☐ | ☐ | ☐ | ☐ |
| I am prepared to make sacrifices to eliminate Covid-19 from New Zealand | ☐ | ☐ | ☐ | ☐ | ☐ |
| Most people are prepared to make sacrifices to eliminate Covid-19 from New Zealand | ☐ | ☐ | ☐ | ☐ | ☐ |
| Most people know we must work together to eliminate Covid-19 from New Zealand | ☐ | ☐ | ☐ | ☐ | ☐ |

**Q4: Involvement with eliminating Covid-19** **from New Zealand**

We are interested in your opinions about eliminating Covid-19 from New Zealand. How strongly do you agree or disagree with the following statements?

| **Item** | **Strongly agree** | **Agree** | **Unsure/**  **neutral** | **Disagree** | **Strongly disagree** |
| --- | --- | --- | --- | --- | --- |
| I think helping to eliminate Covid-19 from New Zealand is rewarding | ☐ | ☐ | ☐ | ☐ | ☐ |
| The consequences are serious if we don’t eliminate Covid-19 from New Zealand | ☐ | ☐ | ☐ | ☐ | ☐ |
| Eliminating Covid-19 from New Zealand is something I am passionate about | ☐ | ☐ | ☐ | ☐ | ☐ |
| It would be a big deal if government made mistakes while we try to eliminate Covid-19 from New Zealand | ☐ | ☐ | ☐ | ☐ | ☐ |
| My position on eliminating Covid-19 from New Zealand tells others something about me | ☐ | ☐ | ☐ | ☐ | ☐ |
| Eliminating Covid-19 from New Zealand is important to me | ☐ | ☐ | ☐ | ☐ | ☐ |
| Making decisions about how to eliminate Covid-19 from New Zealand is complicated | ☐ | ☐ | ☐ | ☐ | ☐ |
| What others think about eliminating Covid-19 from New Zealand tells me something about them | ☐ | ☐ | ☐ | ☐ | ☐ |
| I care a lot about eliminating Covid-19 from New Zealand | ☐ | ☐ | ☐ | ☐ | ☐ |
| Making decisions about how to eliminate Covid-19 from New Zealand is difficult | ☐ | ☐ | ☐ | ☐ | ☐ |

**Q5: Involvement with vaccinating to help eliminate Covid-19?**

The government is hoping to use vaccines to help eliminate Covid-19 from New Zealand. How strongly do you agree or disagree with the following statements about having a vaccine for Covid-19?

| **Item** | **Strongly agree** | **Agree** | **Unsure/**  **neutral** | **Disagree** | **Strongly disagree** |
| --- | --- | --- | --- | --- | --- |
| Getting vaccinated against Covid-19 would be rewarding | ☐ | ☐ | ☐ | ☐ | ☐ |
| The consequences are serious if mistakes are made with getting vaccinated against Covid-19 | ☐ | ☐ | ☐ | ☐ | ☐ |
| Getting vaccinated against Covid-19 is something I am passionate about | ☐ | ☐ | ☐ | ☐ | ☐ |
| It would be a big deal if I made a mistake with getting vaccinated against Covid-19 | ☐ | ☐ | ☐ | ☐ | ☐ |
| My position about getting vaccinated for Covid-19 tells others something about me | ☐ | ☐ | ☐ | ☐ | ☐ |
| Getting vaccinated against Covid-19 is important to me | ☐ | ☐ | ☐ | ☐ | ☐ |
| Making decisions about getting vaccinated against Covid-19 is complicated | ☐ | ☐ | ☐ | ☐ | ☐ |
| What others think about getting vaccinated against Covid-19 tells me something about them | ☐ | ☐ | ☐ | ☐ | ☐ |
| I care a lot about getting vaccinated against Covid-19 | ☐ | ☐ | ☐ | ☐ | ☐ |
| Making decisions about getting vaccinated against Covid-19 is difficult | ☐ | ☐ | ☐ | ☐ | ☐ |

**Q6: Attitude towards getting vaccinated to stop the spread Covid-19**

How strongly do you agree or disagree with the following statements about getting vaccinated to stop the spread of Covid-19?

| **Item** | **Strongly agree** | **Agree** | **Unsure/**  **neutral** | **Disagree** | **Strongly disagree** |
| --- | --- | --- | --- | --- | --- |
| I think people should get vaccinated to help stop the spread of Covid-19 | ☐ | ☐ | ☐ | ☐ | ☐ |
| I think getting vaccinated against Covid-19 is the right thing to do | ☐ | ☐ | ☐ | ☐ | ☐ |
| I believe it is wrong to get vaccinated against Covid-19 | ☐ | ☐ | ☐ | ☐ | ☐ |
| I think it would be good to get vaccinated against Covid-19 | ☐ | ☐ | ☐ | ☐ | ☐ |

**Q7:** Which one of the following statements best describes you?

Please choose one

| **Item** | **Describes me** |
| --- | --- |
| I really think getting vaccinated against Covid-19 is the right thing to do | ☐ |
| It doesn’t really matter to me whether or not I get vaccinated against Covid-19 | ☐ |
| I am not really sure if getting vaccinated against Covid-19 is the best way to go | ☐ |
| I haven’t put much thought into getting vaccinated against Covid-19 | ☐ |
| I strongly believe that getting vaccinated against Covid-19 is a bad thing to do | ☐ |

**Q8: Perceived advantages and disadvantages of getting vaccinated against Covid-19**

How strongly do you agree or disagree with the following statements about getting vaccinated against Covid-19?

| **Item** | **Strongly agree** | **Agree** | **Unsure/**  **neutral** | **Disagree** | **Strongly disagree** |
| --- | --- | --- | --- | --- | --- |
| A vaccine will give lifelong protection against Covid-19 | ☐ | ☐ | ☐ | ☐ | ☐ |
| Getting vaccinated against Covid-19 is just not practical | ☐ | ☐ | ☐ | ☐ | ☐ |
| Getting vaccinated against Covid-19 isn’t worthwhile if you are only protected for a few months | ☐ | ☐ | ☐ | ☐ | ☐ |
| Getting vaccinated against Covid-19 means you will recover faster | ☐ | ☐ | ☐ | ☐ | ☐ |
| Getting vaccinated against Covid-19 should be compulsory | ☐ | ☐ | ☐ | ☐ | ☐ |
| You should only have to get vaccinated if you are old or have a health problem | ☐ | ☐ | ☐ | ☐ | ☐ |
| Getting vaccinated against Covid-19 means your symptoms will be much weaker if you do get the virus | ☐ | ☐ | ☐ | ☐ | ☐ |
| Getting vaccinated is a waste of time and effort | ☐ | ☐ | ☐ | ☐ | ☐ |
| It isn’t worth getting vaccinated yet as there are too many unknowns about the vaccines | ☐ | ☐ | ☐ | ☐ | ☐ |
| Vaccination against Covid-19 should be free | ☐ | ☐ | ☐ | ☐ | ☐ |
| Children shouldn’t have to be vaccinated against Covid-19 | ☐ | ☐ | ☐ | ☐ | ☐ |
| People who want to be vaccinated against Covid-19 are over-reacting | ☐ | ☐ | ☐ | ☐ | ☐ |
| Once you are vaccinated you cannot catch Covid-19 | ☐ | ☐ | ☐ | ☐ | ☐ |
| I think we should wait and see if vaccination works overseas before trying it here | ☐ | ☐ | ☐ | ☐ | ☐ |
| Getting vaccinated against Covid-19 is unsafe because of the potential side effects | ☐ | ☐ | ☐ | ☐ | ☐ |
| Once you are vaccinated you cannot spread Covid-19 | ☐ | ☐ | ☐ | ☐ | ☐ |

**Q9: Once a vaccine for Covid-19 is available, will you get vaccinated? Please choose one.**

| ☐ | Definitely | Go to Q10 |
| --- | --- | --- |
| ☐ | Probably | Go to Q10 |
| ☐ | Maybe | Go to Q12 |
| ☐ | Probably not | Go to Q12 |
| ☐ | Definitely not | Go to Q12 |

**Q10: Once a vaccine is available would you get vaccinated as soon as you could?**

| ☐ | Yes | Go to Q11 |
| --- | --- | --- |
| ☐ | Not sure | Go to Q11 |
| ☐ | No | Go to Q12 |

**Q11: Would you get vaccinated if the vaccine only offered protection for a few months?**

| ☐ | Yes |
| --- | --- |
| ☐ | Not sure |
| ☐ | No |

**Q12: Have you had a bad experience with a vaccination?**

| ☐ | Yes |
| --- | --- |
| ☐ | Not sure |
| ☐ | No |

**Q13: Do you know someone who had a bad experience with a vaccination?**

| ☐ | Yes |
| --- | --- |
| ☐ | Not sure |
| ☐ | No |

**Part B: Demographics**

We just have a few questions to make sure we get a good cross-section of people.

#### B1: What age bracket do you fit into?

( ) 18-29 years

( ) 30-39 years

( ) 40-49 years

( ) 50-59 years

( ) 60-69 years

( ) 70 years and over

( ) Prefer not to say

#### B2: Which of the following do you identify as?

( ) Male

( ) Female

( ) Gender diverse

( ) Prefer not to say

#### B3: What is your highest level of formal education?

( ) Some or all of secondary school

( ) Certificate (1-6)

( ) Diploma (5-7)

( ) Bachelor degree

( ) Post-graduate diploma/certificate

( ) Post-graduate degree

( ) Prefer not to say

#### B4: Which of the following do you identify with?

( ) Māori

( ) European New Zealander

( ) Pacific Islander

( ) Asian

( ) Other

( ) Prefer not to say

#### B5: What household income bracket do you fit into?

( ) Less than $20,000

( ) $20,000 to $50,000

( )$50,000 to $70,000

( ) $70,000 to $100,000

( ) more than $100,000

( ) Prefer not to say

**Thank you so much for participating in our survey.**
