## Appendix A for "Predicting willingness to be vaccinated for Covid-19: evidence from New Zealand"

Appendix A – Sample demographics

Table A1. Age distribution of respondents

| Age category (years) | Percentage of respondents | Percentage of New Zealand residents^1^ |
| --- | --- | --- |
| 18–29 | 16.3 | 19.6 |
| 30–39 | 19.4 | 17.8 |
| 40–49 | 19.1 | 17.4 |
| 50–59 | 15.7 | 17.5 |
| 60–69 | 14.4 | 13.9 |
| 70 and over | 14.8 | 13.7 |

Notes:	^1^Source: [50]

Table A2. Distribution of respondents by highest educational qualification

| Education category | Percentage of respondents | Percentage of New Zealand residents^1^ |
| --- | --- | --- |
| Some or all of secondary school | 21.6 | 19.3 |
| Certificate (1–6) | 17.3 | 43.8 |
| Diploma (5–7) | 16.1 | 10.4 |
| Graduate or postgraduate | 45.0 | 26.4 |

Notes:	^1^Source: [51]

Table A3. Ethnicity distribution of respondents

| Ethnic category | Percentage of respondents | Percentage of New Zealand residents^1^ |
| --- | --- | --- |
| European | 70.9 | 62.4 |
| Māori | 5.7 | 14.8 |
| Pacific Islander | 2.6 | 7.4 |
| Asian | 12.0 | 14.0 |
| Other | 7.6 | 1.4 |

Notes:	^1^Source: [50]

Table A4. Income distribution of respondents

| Income category | Percentage of respondents | Approximate percentage of New Zealand households^1^ |
| --- | --- | --- |
| Less than $20,000 | 6.4 | 10.0 |
| $20,000 to $50,000 | 25.6 | 20.0 |
| $50,000 to $70,000 | 15.8 | 20.0 |
| More than $70,000 | 40.3 | 50.0 |

Notes:	^1^ Based on household income deciles. First decile <$25,400, second and third deciles $25,400 to $52,199, fourth and fifth deciles %52,200 to $82,999, remaining deciles >$83,000. Source: [52]
