## Appendix B for "Predicting willingness to be vaccinated for Covid-19: evidence from New Zealand"

**Appendix B: Belief segments results**

**Table B1.** Belief segments for Covid-19

| **Statement** | **Covid-19 convinced (37%)** | **Covid-19 moderates (40%)** | **Covid-19 ambivalents (12%)** | **Covid-19 asymptomatics**  **(5%)** | **Covid-19 sceptics**  **(6%)** |
| --- | --- | --- | --- | --- | --- |
| Coughing and sneezing spreads Covid-19 | 4.65 | 4.30 ^a^ | 3.64 ^a, b^ | 2.77 ^a, b, c^ | 4.25 ^a, c, d^ |
| Covid-19 spreads from surfaces touched by infected people | 4.35 | 3.84 ^a^ | 3.25 ^a, b^ | 2.06 ^a, b, c^ | 4.11 ^c, d^ |
| Covid-19 is only a danger to the elderly and people with health problems | 1.32 | 2.21 ^a^ | 3.61 ^a, b^ | 1.48 ^b, c^ | 4.25 ^a, b, c^ |
| You are immune to re-infection once you have had Covid-19 | 1.95 | 2.47 ^a^ | 2.86 ^a, b^ | 1.77 ^b, c^ | 4.07 ^a, b, c, d^ |
| Children cannot catch Covid-19 | 1.23 | 1.72 ^a^ | 2.59 ^a, b^ | 1.29 ^b, c^ | 3.75 ^a, b, c, d^ |
| Children are perfectly safe from Covid-19 | 1.25 | 1.81 ^a^ | 2.81 ^a, b^ | 1.38 ^b, c^ | 4.16 ^a, b, c, d^ |
| You cannot catch the virus from people without symptoms | 1.27 | 2.09 ^a^ | 2.90 ^a, b^ | 1.40 ^b, c^ | 3.98 ^a, b, c, d^ |
| Covid-19 is a hoax | 1.11 | 1.53 ^a^ | 2.61 ^a, b^ | 1.21 ^b, c^ | 3.97 ^a, b, c, d^ |
| Fears about Covid-19 are exaggerated | 1.45 | 2.45 ^a^ | 3.48 ^a, b^ | 1.56 ^b, c^ | 4.41 ^a, b, c, d^ |
| Covid-19 is no worse than the seasonal flu | 1.33 | 2.23 ^a^ | 3.09 ^a, b^ | 1.48 ^b, c^ | 4.07 ^a, b, d^ |
| Covid-19 is man-made | 2.23 | 2.70 ^a^ | 3.19 ^a, b^ | 2.27 ^b, c^ | 4.00^a, b, c, d^ |
| Covid-19 comes from bats | 2.96 | 2.79 | 2.67 ^a^ | 2.40 | 4.10 ^a, b, c, d^ |

Notes:	Values are mean agreement ratings. Ratings ranged from a minimum of 1 (strongly disagree) to a maximum of 5 (strongly agree).

Differences in mean agreement ratings between segments tested using Tukey’s HSD [43], (p<0.01)

^a^ Mean differs from mean for the ‘convinced’ segment

^b^ Mean differs from mean for the ‘moderates’ segment

^c^ Mean differs from the mean for the ‘ambivalents’ segment

^d^ Mean differs from the mean for the ‘asymptomatics’ segment

**Table B2. Belief segments for eliminating Covid-19**

| **Statement** | **Elimination enthusiasts (26%)** | **Elimination moderates (18%)** | **Vaccine**  **hopefuls (34%)** | **Elimination sceptics (22%)** |
| --- | --- | --- | --- | --- |
| We need to eliminate Covid-19 to save lives | 4.67 | 3.53 ^a^ | 4.37 ^a, b^ | 3.46 ^a, c^ |
| We should just live with it until we have a vaccine | 1.44 | 2.72 ^a^ | 2.63 ^a,^ | 3.83 ^a, b, c^ |
| It would be better to let it spread and build herd immunity | 1.26 | 2.19 ^a^ | 1.69 ^a, b^ | 3.70 ^a, b, c^ |
| There is no point trying to eliminate Covid-19 because it is a virus and will keep changing | 1.66 | 2.49 ^a^ | 2.76 ^a, b^ | 3.99 ^a, b, c^ |
| Covid-19 is everywhere in the world so there is no way we can keep it out | 1.71 | 2.26 ^a^ | 3.80 ^a, b^ | 4.16 ^a, b, c^ |

Notes:	Values are mean agreement ratings. Ratings ranged from a minimum of 1 (strongly disagree) to a maximum of 5 (strongly agree).

Differences in mean agreement ratings between segments tested using Tukey’s HSD test [43] (p<0.01)

^a^ Mean differs from mean for the ‘enthusiasts’ segment

^b^ Mean differs from mean for the ‘moderates’ segment

^c^ Mean differs from the mean for the ‘hopefuls’ segment

**Table B3. Belief segments for Covid-19 vaccination**

| **Statement** | **Vaccination enthusiasts**  **(24%)** | **Vaccination moderates**  **(33%)** | **Vaccination ambivalent**  **(28%)** | **Vaccination cautious**  **(7%)** | **Vaccination sceptics**  **(8%)** |
| --- | --- | --- | --- | --- | --- |
| A vaccine will give lifelong protection against Covid-19 | 2.46 | 2.76 ^a^ | 2.76 ^a^ | 4.04 ^a, b, c^ | 1.46 ^a, b, c, d^ |
| Getting vaccinated against Covid-19 means you will recover faster | 3.58 | 3.39 ^a^ | 2.98 ^a, b^ | 4.08 ^a, b, c^ | 2.14 ^a, b, c, d^ |
| Getting vaccinated against Covid-19 means your symptoms will be much weaker if you do get the virus | 3.76 | 3.48 ^a^ | 3.25 ^a, b^ | 4.11 ^a, b, c^ | 2.63 ^a, b, c, d^ |
| Once you are vaccinated you cannot catch Covid-19 | 2.51 | 2.74 ^a^ | 2.63 | 4.19 ^a, b, c^ | 1.60 ^a, b, c, d^ |
| Once you are vaccinated you cannot spread Covid-19 | 2.75 | 3.02 ^a^ | 2.87 | 3.95 ^a, b, c^ | 1.68 ^a, b, c, d^ |
| You should only have to get vaccinated against Covid-19 if you are old or have a health problem | 1.16 | 2.02 ^a^ | 2.76 ^a, b^ | 4.05 ^a, b, c^ | 2.79 ^a, b, d^ |
| Children shouldn't be vaccinated against Covid-19 | 1.70 | 2.25 ^a^ | 2.94 ^a, b^ | 3.87 ^a, b, c^ | 3.71 ^a, b, c^ |
| Getting vaccinated against Covid-19 is a waste of time and effort | 1.07 | 1.69 ^a^ | 2.62 ^a, b^ | 3.89 ^a, b, c^ | 3.46 ^a, b, c, d^ |
| People who want to be vaccinated against Covid-19 are over-reacting | 1.17 | 2.09 ^a^ | 2.66 ^a, b^ | 4.11 ^a, b, c^ | 3.09^a, b, c, d^ |

Notes:	Values are mean agreement ratings. Ratings ranged from a minimum of 1 (strongly disagree) to a maximum of 5 (strongly agree).

Differences in mean agreement ratings between segments tested using Tukey’s HSD [43], (p<0.01)

^a^ Mean differs from mean for the ‘enthusiasts’ segment

^b^ Mean differs from mean for the ‘moderates’ segment

^c^ Mean differs from the mean for the ‘ambivalents’ segment

^d^ Mean differs from the mean for the ‘cautious’ segment

**Table B3. Belief segments for Covid-19 vaccination (continued)**

| **Statement** | **Vaccination enthusiasts**  **(24%)** | **Vaccination moderates (33%)** | **Vaccination ambivalent**  **(28%)** | **Vaccination cautious**  **(7%)** | **Vaccination sceptics**  **(8%)** |
| --- | --- | --- | --- | --- | --- |
| Getting vaccinated against Covid-19 should be compulsory | 3.97 | 3.57 ^a^ | 2.54 ^a, b^ | 4.05 ^b, c^ | 2.79 ^a, b, d^ |
| Vaccination against Covid-19 should be free | 4.81 | 4.44 ^a^ | 4.04 ^a, b^ | 4.23 ^a^ | 4.10^a, b^ |
| It isn't worth getting vaccinated against Covid-19 yet as there are too many unknowns about the vaccines | 1.43 | 2.35 ^a^ | 3.41 ^a, b^ | 4.27 ^a, b, c^ | 4.53 ^a, b, c^ |
| I think we should wait and see if the Covid-19 vaccination works overseas before trying it here | 1.75 | 2.59 ^a^ | 3.70 ^a, b^ | 3.93 ^a, b^ | 4.19 ^a, b, c^ |
| Getting vaccinated against Covid-19 is unsafe because of the potential side effects | 1.46 | 2.29 ^a^ | 3.22 ^a, b^ | 3.96 ^a, b, c^ | 4.10 ^a, b, c^ |
| Getting vaccinated against Covid-19 is just not practical | 1.15 | 1.95 ^a^ | 2.71 ^a, b^ | 4.13 ^a, b, c^ | 3.26 ^a, b, c, d^ |
| Getting vaccinated against Covid-19 isn't worthwhile if you are only protected for a few months | 1.70 | 2.52 ^a^ | 3.44 ^a, b^ | 4.07 ^a, b, c^ | 4.09 ^a, b, c^ |

Notes:	Values are mean agreement ratings. Ratings ranged from a minimum of 1 (strongly disagree) to a maximum of 5 (strongly agree).

Differences in mean agreement ratings between segments tested using Tukey’s HSD [43], (p<0.01)

^a^ Mean differs from mean for the ‘enthusiasts’ segment

^b^ Mean differs from mean for the ‘moderates’ segment

^c^ Mean differs from the mean for the ‘ambivalents’ segment

^d^ Mean differs from the mean for the ‘cautious’ segmen

**Table B4. Vaccination belief segments and attitude towards being vaccinated**

| **Segment** | **Right thing to do** | **Doesn’t matter to me** | **Not sure** | **Haven’t given it much thought** | **Bad thing to do** |
| --- | --- | --- | --- | --- | --- |
| Vaccination enthusiasts | 99.6 | 0.0 | 0.0 | 0.4 | 0.0 |
| Vaccination moderates | 90.9 | 4.0 | 4.8 | 1.2 | 0.0 |
| Vaccination cautious | 54.7 | 21.3 | 12.0 | 5.3 | 6.7 |
| Vaccination ambivalent | 33.6 | 9.6 | 47.1 | 7.5 | 2.1 |
| Vaccination sceptics | 0.0 | 6.3 | 48.8 | 7.5 | 37.5 |

Notes:	Values are percentages of respondents in each segment. Test for differences in percentages across segments (χ^2^ = 726.2, *P* < 0.01)

**Table B5. Vaccination belief segments and willingness to be vaccinated**

| **Segment** | **Definitely** | **Probably** | **Maybe** | **Probably not** | **Definitely not** |
| --- | --- | --- | --- | --- | --- |
| Vaccination enthusiasts | 92.4 | 7.2 | 0.4 | 0.0 | 0.0 |
| Vaccination moderates | 66.7 | 27.0 | 5.2 | 1.2 | 0.0 |
| Vaccination cautious | 44.0 | 32.0 | 13.3 | 9.3 | 1.0 |
| Vaccination ambivalent | 13.6 | 21.8 | 46.4 | 13.2 | 5.0 |
| Vaccination sceptics | 0.0 | 0.0 | 18.8 | 26.3 | 55.0 |

Notes:	Values are percentages of respondents in each segment. Test for differences in percentages across segments (χ^2^ = 427.8, *P* < 0.01).

**Table B6. Vaccination belief segments and willingness to be vaccinated as soon as possible**

| **Segment** | **Yes** | **Not sure** | **No** |
| --- | --- | --- | --- |
| Vaccination enthusiasts | 91.1 | 5.5 | 3.4 |
| Vaccination moderates | 76.7 | 18.1 | 5.2 |
| Vaccination cautious | 82.1 | 6.0 | 11.9 |
| Vaccination ambivalent | 23.1 | 49.3 | 27.5 |
| Vaccination sceptics | 0.0 | 26.7 | 73.3 |

Notes:	Values are percentages of respondents in each segment who indicated they definitely, probably or maybe would get vaccinated for Covid-19.

Test for differences in percentages across segments (χ^2^ = 339.8, *P* < 0.01).

**Table B7. Vaccination belief segments and willingness to be vaccinated if it offers only temporary protection**

| **Segment** | **Yes** | **Not sure** | **No** |
| --- | --- | --- | --- |
| Vaccination enthusiasts | 82.5 | 13.5 | 3.9 |
| Vaccination moderates | 60.5 | 28.2 | 11.3 |
| Vaccination cautious | 86.4 | 5.1 | 8.5 |
| Vaccination ambivalent | 19.3 | 48.8 | 31.9 |
| Vaccination sceptics | 0.0 | 50.0 | 50.0 |

Notes:	Values are percentages of respondents in each segment who indicated they would get vaccinated as soon as possible for Covid-19.

Test for differences in percentages across segments (χ2 = 194.7, P < 0.01).

**Table B8. Vaccination belief segments and bad experience with vaccinations**

| **Segment** | **Yes** | **Not sure** | **No** |
| --- | --- | --- | --- |
| Vaccination enthusiasts | 5.1 | 2.1 | 92.8 |
| Vaccination moderates | 7.9 | 2.1 | 90.0 |
| Vaccination cautious | 42.7 | 8.2 | 56.0 |
| Vaccination ambivalent | 13.6 | 3.9 | 78.2 |
| Vaccination sceptics | 32.5 | 1.3 | 66.3 |

Notes:	Values are percentages of respondents in each segment.

Test for differences in percentages across segments (χ2 = 126.2, P < 0.01).

**Table B9. Vaccination belief segments and knowing someone who had a bad experience with vaccinations**

| **Segment** | **Yes** | **Not sure** | **No** |
| --- | --- | --- | --- |
| Vaccination enthusiasts | 15.6 | 4.2 | 80.2 |
| Vaccination moderates | 17.9 | 3.6 | 78.5 |
| Vaccination cautious | 46.7 | 2.7 | 50.7 |
| Vaccination ambivalent | 25.0 | 11.8 | 63.2 |
| Vaccination sceptics | 24.6 | 6.3 | 69.2 |

Notes:	Values are percentages of respondents in each segment.

Test for differences in percentages across segments (χ^2^ = 107.9, *P* < 0.01)
